# A hospital-wide response to multiple outbreaks of COVID-19 in Health Care Workers Lessons learned from the field

**DOI:** 10.1101/2020.09.02.20186452

**Authors:** Kirsty Buising, Deborah Williamson, Benjamin Cowie, Jennifer MacLachlan, Liz Orr, Chris MacIsaac, Eloise Williams, Katherine Bond, Stephen Muhi, James McCarthy, Andrea B. Maier, Louis Irving, Denise Heinjus, Cate Kelly, Caroline Marshall

## Abstract

**Objective:** To describe COVID-19 infections amongst healthcare workers at the Royal Melbourne Hospital from 1^st^ July to 31^st^ August 2020

**Design:** Prospective observational study

**Setting:** A 550 bed tertiary referral hospital in metropolitan Melbourne

**Participants:** All healthcare workers identified with COVID-19 infection in the period of interest

**Results:** 262 healthcare worker infections were identified over 9 weeks. 68.3% of infected healthcare workers were nurses and the most affected locations were the geriatric and rehabilitation wards. Clusters of infection occurred in staff working in wards with patients known to have COVID-19 infection. Staff infections peaked when COVID-19 infected inpatient numbers were highest, and density of patients and certain patient behaviours were noted by staff to be linked to possible transmission events. Three small outbreaks on other wards occurred but all were recognised and brought under control. Availability of rapid turn-around staff testing, and regular review of local data and obtaining feedback from staff helped identify useful interventions which were iteratively implemented. Attention to staff wellbeing was critical to the response and a comprehensive support service was implemented.

**Conclusion(s):** A comprehensive multimodal approach to containment was instituted with iterative refinement based on frontline workers observations and ongoing analysis of local data in real time.

The known: Healthcare workers are a group recognized to be at risk of acquisition of infection in the workplace during the current COVID-19 pandemic
The new: This describes the experience of the largest Australian outbreak to date of COVID- 19 infection amongst healthcare workers in a hospital environment
The implications: This paper should assist healthcare services to prepare for surges in COVID-19 infection to help limit future transmissions to healthcare workers

## INTRODUCTION

The COVID-19 (the illness caused by novel coronavirus) pandemic represents a global crisis. At September 2^nd^ 2020, there have been more than 25 million cases of COVID-19 reported to the World Health Organization, with over 800,000 deaths globally (1). In many countries, high rates of community infection have resulted in extreme pressure on health services, with an observed increased risk of COVID-19 infection in front-line healthcare workers. (2) To date, factors associated with COVID-19 infection in healthcare workers overseas have included inadequate personal protective equipment (PPE), exposure to large numbers of patients with COVID-19, worker fatigue, and limited access to diagnostic testing leading to potential under-recognition of cases (2–4).

In Australia, the prevalence of COVID-19 remains comparatively low compared to many other countries. A ‘first wave’ of COVID-19 occurred in March / April, prompting implementation of nationwide public health interventions, including closing of the Australian international border (5). During this initial phase of the epidemic, infections in healthcare workers were largely attributable to transmission events arising from returning international travellers, as evidenced by distinct lineages of SARS-CoV-2 (Severe Acute Respiratory Virus Coronavirus 2) the causative agent of COVID-19 circulating in infected patients (5). This observation was also made amongst infected healthcare workers at our own health service (6).

Since mid-June, the state of Victoria has experienced a surge in COVID-19 cases requiring re-implementation of a range of public health interventions. Concurrent with this, there was a significant increase in COVID-19 infection amongst staff. At our institution, the Royal Melbourne Hospital (RMH) we observed a marked increase in infections amongst hospital staff during July and August 2020. Although considerable planning had been undertaken in preparation for possible outbreaks in our setting, we identified a number of ‘real-world’ challenges in controlling staff infections, particularly the sheer rapidity of disease spread. To inform future responses to healthcare worker infections in the Australian setting, we present a descriptive epidemiological analysis of healthcare worker infections at our institution and describe the suite of interventions associated with outbreak control.

## METHODS

### Study setting

The RMH is a 550-bed University-affiliated tertiary hospital in metropolitan Melbourne, Australia. The hospital manages a state-wide trauma service, state-wide allogeneic stem cell transplant service, as well as providing renal transplantation, neurosurgical and cardiac surgical services. In addition, the RMH manages geriatric and rehabilitation services at its Royal Park Campus (RPC); the state’s largest mental health service, North West Mental Health (NWMH); and four residential aged care homes. Overall there are approximately 10,000 staff employed by RMH. These employees (which are all included in the term HCWs) include clinical staff (nursing, medical, allied health), support staff (including cleaners, food services, security), technical staff (e.g. laboratory personnel), and administrative staff.

### Management of symptomatic HCW

The health service provided a rapid access clinic for symptomatic staff to obtain COVID-19 testing throughout the pandemic. In addition all people tested in the public screening clinic were asked to identify themselves as healthcare workers. Concurrently, all positive tests state-wide were reported to the Victorian Department of Health and Human Services (DHHS), and infections in healthcare workers were reported back to their health service (to capture anyone tested at outside laboratories). Every infected HCW had a detailed interview conducted by a trained infection prevention clinical nurse consultant /contact tracer to identify close contacts amongst the staff, patients or visitors. The affected staff member’s use of PPE was described in detail as well as the locations of work in the days prior to infection, nature of their work, types of patients cared for, and any suspected acquisition events nominated by the healthcare worker. Contacts with other staff before and after work and at breaks were explored, as well as possible community contacts. Staff with COVID-19 were required to isolate at home or in a hotel for at least 10 days post symptom onset, according to DHHS guidelines. Staff contacts were furloughed from the workplace for 14 days and quarantined. These were typically ‘close’ contacts as defined by DHHS guidelines. Patient contacts were managed using appropriate infection control precautions, and community contacts were advised to home quarantine. Outbreaks, defined as two or more epidemiologically and / or spatially linked cases in healthcare workers and/or patients, were managed by a multidisciplinary incident management team. Records of decisions made were collected prospectively in minutes from meetings held daily by the Emergency Operations Centre (EOC) which was activated under RMH Emergency plan to provide central command for the health service.

### Testing of asymptomatic HCW

Health service-wide asymptomatic staff testing was offered voluntarily in May 2020 (prior to this study’s inclusion period following public health instruction) and again from 25th to 31st July 2020 (initiated by the hospital, again voluntary). In addition, in the context of focal ward outbreaks, targeted asymptomatic screening occurred for staff and patients in affected wards, initially weekly and later twice weekly.

### Microbiological testing

Deep nasal and throat swabs were used to obtain samples for nucleic acid testing (Reverse Transcription-Polymerase Chain Reaction for SARS-CoV2). SARS-CoV-2 RNA was detected using the Coronavirus Typing assay (AusDiagnostics), a 2-step, hemi-nested multiplex tandem PCR, with 7 coronavirus RNA targets plus a proprietary artificial sequence as an internal control (7). All positive samples were subject to additional confirmatory PCR testing for SARS-CoV-2 at a local reference laboratory, using previously published primers (8).

### Statistical analysis

Data regarding HCW infections were entered into data collection project in REDCap (v10), then extracted into an Excel spreadsheet and analysed in Stata (v16).

### Ethics

Data reported in this study were collected as part of the usual infection prevention service quality assurance activities. This study was approved by the Melbourne Health Human Research Ethics Committee as QA2020058.

## RESULTS

### Overview of HCW infections

Over a nine week period between 1st July 2020 and 31st August 2020, 262 cases of COVID-19 were identified amongst RMH staff (Figure 1). Of cases, 21.7% were in males and 78.3% were in females, with a median age of 32.7 years (interquartile range 26.8 to 44.9 years) (Table 1). Overall, 28 HCWs (10.6%) with COVID-19 infection needed hospitalization, including 13 who required admission to inpatient wards, while 15 required a hospital-in-the-home admission (inpatient bed-substitution care in a patient’s own home). Two staff required admission to the intensive care unit (ICU), none required mechanical ventilation and there were no deaths. Of those whose roles were known, nurses were the most affected staff group comprising 68.3% followed by support staff (such as food and cleaning services staff) 14.5% and then doctors 8.0% (see Table 1). Of the affected medical staff, 17/21 were doctors-in-training. The majority of staff infections occurred in staff working on geriatric and rehabilitation inpatient wards at RPC, and in acute wards with large numbers of COVID-19 positive patients managed concurrently (“hot wards”). No operating theatre staff and no staff working in the four residential aged care facilities managed by RMH were identified as being COVID-19 positive in the time period described. The ICU had between 0 and 10 (median 7) concurrent patients with COVID-19 infection over the period, and 4 staff in the ICU were identified as having acquired COVID-19 infection. The median turnaround time for healthcare worker COVID-19 test results (from specimen collection to reporting) was 20.2 hours (interquartile range 11.4 to 29.1 hours). Overt recognized PPE breeches were rarely reported. Contacts with known COVID-19 cases outside the hospital were infrequent, but on occasion did occur (e.g, two affected healthcare workers who lived together). Overall 25 % of the staff who were furloughed ultimately tested positive for COVID-19.

**Figure 1.**
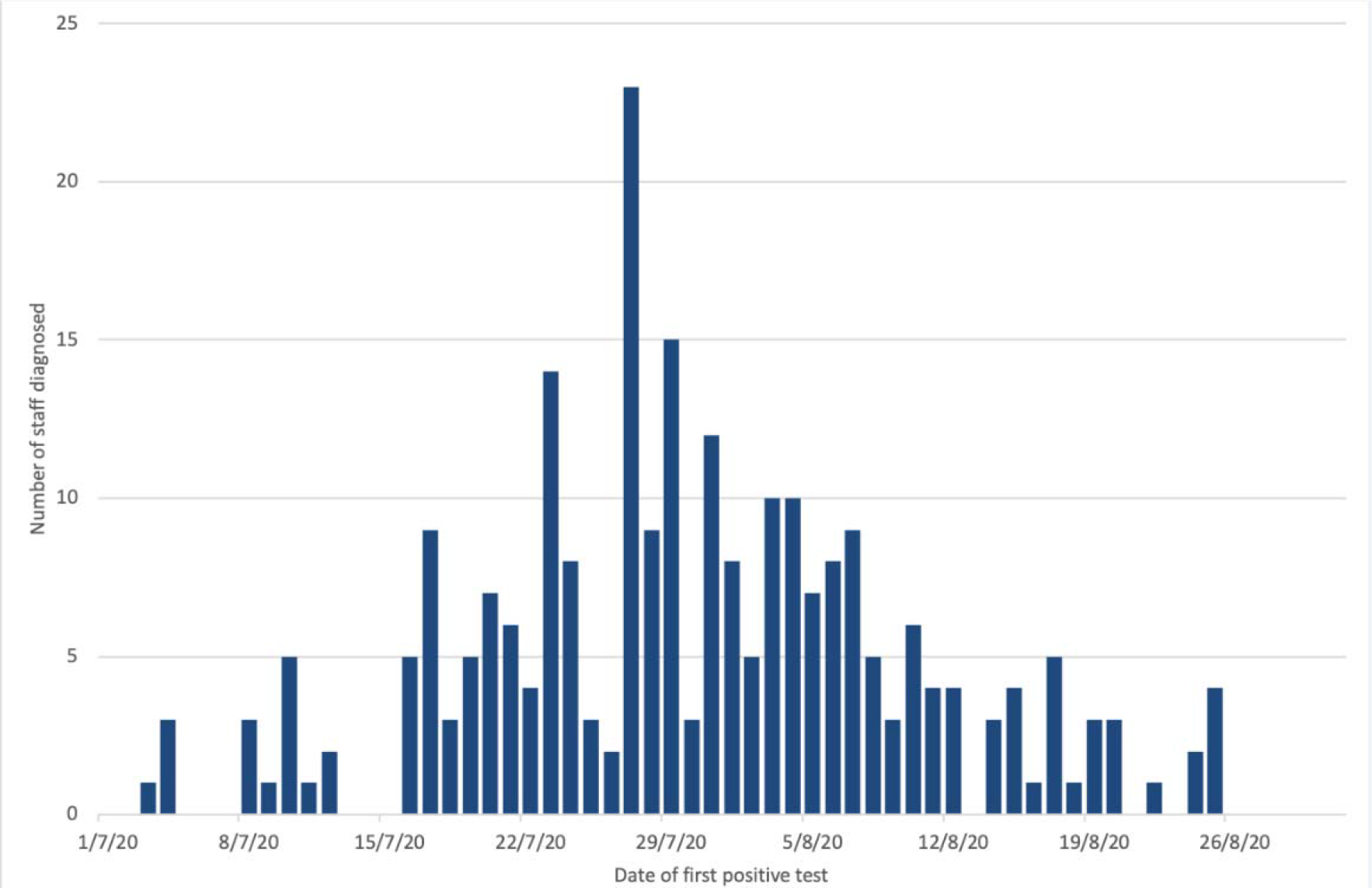
Epidemic curve of healthcare worker infections at the Royal Melbourne Hospital, 1^st^ July 2020 to 31^st^ August 2020.

**Table 1.** Demographic characteristics of healthcare workers with PCR-confirmed COVID-19 infection at the Royal Melbourne Hospital, July 1^st^ to August 31st, 2020.

| Characteristic | Number (% of total positives)<br>N=262 |
| --- | --- |
| Sex: |  |
| Male | 57 (21.8) |
| Female | 205 (78.2) |
| Median age at diagnosis (IQR) | 32.7 (26.8 to 44.9) years |
| Employee type: |  |
| Nurse | 179 (68.3) |
| Doctor | 21 (8.0) |
| Allied Health | 9 (3.4) |
| Support staff (food services; environmental services) | 38 (14.5) |
| Administrative staff | 6 (2.3) |
| Students | 4 (1.5) |
| Security staff | 4 (1.5) |
| Laboratory staff | 1 (0.4) |
| Location: |  |
| RPC | 107 (40.8) |
| Hot wards (COVID wards, ED, ICU) | 57 (21.8) |
| Cold wards with recognized COVID-19 outbreaks (3 wards) | 20 (7.6) |
| Cold wards with no outbreaks (1 or 2 unlinked cases - 6 wards) | 7 (2.7) |
| Mental health ward | 8 (3.1) |
| Not ward based (e.g.; non clinical) | 31 (11.8) |
| Unknown (no campus/ward stated) | 32 (12.2) |

RPC=Royal Park Campus (rehabilitation, geriatric rehabilitation), ED = emergency department, ICU = intensive care unit, COVID wards = known or suspected COVID-19 positive patients managed. Hot ward = ward dedicated to managing confirmed or suspected COVID-19 positive patients. Cold ward = all other wards

### Outbreak linked geriatric and geriatric rehabilitation inpatient wards

The highest number of COVID-19 infected inpatients was at RPC; overall, RPC staff constituted approximately 10% of the total staff workforce (the exact number is difficult to ascertain as some staff move between sites), but RPC staff made up 40.8% (n=107) of COVID-19 healthcare worker infections. Between 12^th^ and 18^th^ July, RPC received a large number of patient admissions from residential aged care facilities with outbreaks of COVID-19. These admitted residents were positive at admission and were managed in transmission-based precautions throughout their stay. PPE was always readily available, and staff were trained in its use. COVID-19 cases in staff rapidly escalated across all six wards at the campus after 16^th^ July, and peaked on 27^th^ July. As new staff infections were recognized, all six wards were progressively moved to being managed with all patients in transmission-based precautions. The peak number of patients with COVID-19 infection being cared for at RPC was 60 patients.

At RPC there are a variety of buildings built from the 1970s to early 2000; most have central air conditioning plants but one area has a local split system. An engineering review of the wards at RPC revealed that air exchanges met current requirements, however, more detailed assessment of air movement was carried out which suggested that some wards were not as well ventilated as others. Some patients were in single rooms, but many were in multi-bed spaces where it was difficult to physically distance patients. Staff reported patients often suffering from a delirium with accompanying wandering behaviours and repeated vocalisation. Nurse specials were often employed to manage these patients. As contact tracing led to large numbers of staff being furloughed, the remaining staff experienced high workloads and care became increasingly difficult. A decision was made by the EOC on 3^rd^ August 2020 to close 4 wards at RPC, a process that was completed by 8^th^ August 2020. Fifteen patients were moved to other health services, while the remaining 45 patients were managed in wards with more modern infrastructure where increased physical separation with patients each having their own room was possible. The closure of these wards coincided with a reduction in HCW infections, although multiple interventions had also been implemented concurrently (Table 2).

**Table 2.**
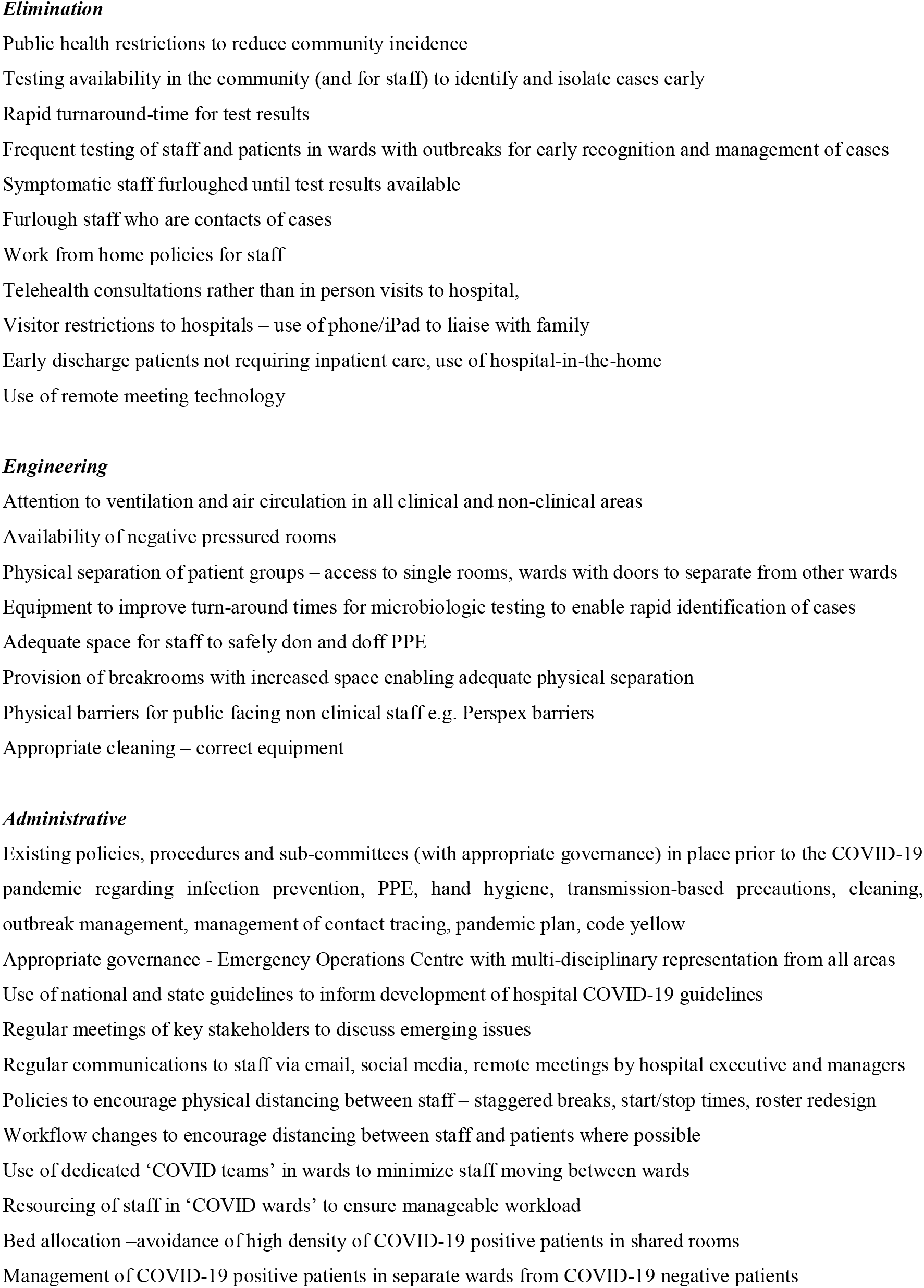
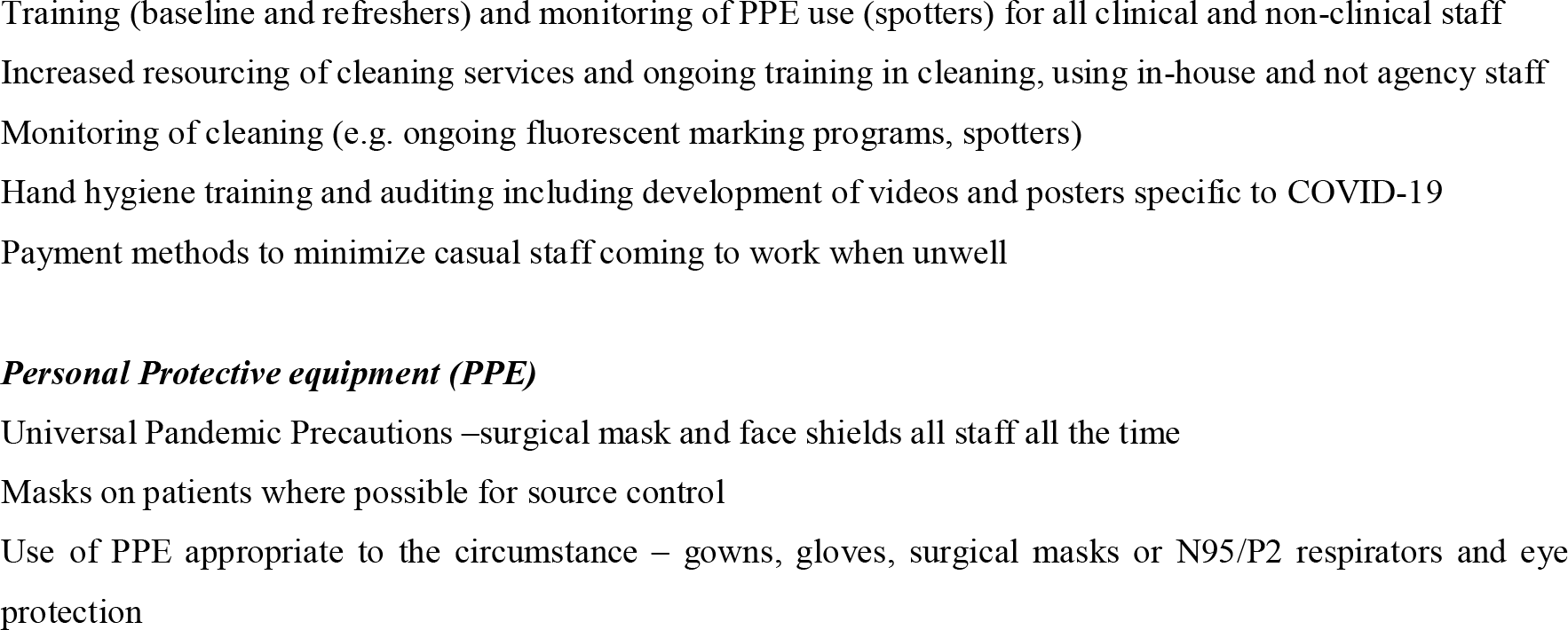
The hierarchy of controls used to guide interventions to address healthcare worker infection with COVID-19 at Royal Melbourne Hospital

### Outbreaks linked to ‘hot wards’ – wards with known /suspected COVID-19 positive patients

At RMH city campus, the majority of affected staff were working in wards with suspected or confirmed COVID-19 infected patients (Table 1). These staff were highly trained in PPE use, appropriate PPE was always readily available, and staff used a ‘buddy’ to check PPE before entry to patient rooms. On one occasion, staff congregation in a tearoom was identified as a likely opportunity for transmission between staff. On other occasions, staff noted that particular behaviours in infected patients appeared to be linked to transmission events (distressed patients shouting, vigorous coughing). The peak combined prevalence of inpatients at RPC and city campus was 99 inpatients on 5^th^ August 2020. Action was undertaken to reduce the density of patients on the COVID-19 wards by ensuring that all patients with COVID-19 had single rooms, where possible. Between 21 and 25^th^ July 2020, the use of N95 particulate respirators by all staff at all times on COVID-19 wards at RMH and RPC was implemented. In addition, ‘spotters’ were deployed to observe PPE donning and doffing; ward walk arounds by senior staff were undertaken regularly to identify emerging issues; wards were de-cluttered; and increased cleaning and active cleaning monitoring were also undertaken. Staff working on hot wards were advised to attend for asymptomatic testing at least weekly when cases of healthcare worker infection had occurred on a ward. The incidence of cases in healthcare workers fell progressively thereafter.

### Outbreaks on ‘cold wards’- wards without known or suspected COVID-19 positive patients

On three occasions, cases in healthcare workers working outside the designated ‘hot wards’ were identified. In some cases, prior work at RPC was identified as a possible risk factor for infection in the index case. The health service developed a management plan for these wards that included: closing the ward to new admissions; moving patients to single rooms (where possible); managing the whole ward using transmission-based precautions; notifying all discharged patients to quarantine in their homes or in the next facility for 14 days; deep cleaning of wards, and testing all patients and staff on each ward at least weekly until no new cases were identified. This testing was voluntary and did identify additional asymptomatic or minimally symptomatic cases. In each cluster at least one additional staff and/or patient case was identified through the enhanced testing. In the context of these staff cases on ‘cold wards’, hospital-wide asymptomatic staff testing occurred from 25th-31st July 2020. This was accompanied by testing of all admitted patients over one week. Over 600 staff and all hospital inpatients were tested with no additional cases identified.

### Institutional responses

The health service responses were multifactorial and iterative with daily review of emerging information that informed ongoing decisions. Case numbers were reviewed daily and reported to the EOC. A dashboard on the hospital intranet was set up to give real time numbers of infected and furloughed staff. Importantly, a ‘hierarchy of controls’ was used to manage these outbreaks, with the importance of each of the measures varying depending on the context of each of the clusters (Table 2). A proactive approach was used to support the wellbeing of both infected and furloughed staff with dedicated nursing and medical staff to monitor physical health (symptoms assessment, oximetry), mental health (counselling) and provide practical supports (delivery of food parcels, blankets, hotel rooms). This service managed over 680 staff during this period.

## DISCUSSION

We describe the largest institutional outbreak of healthcare worker infections with SARSCoV-2 reported in Australia to date. Management of outbreaks is core business for infection prevention services and is based on an established and effective suite of evidence-based interventions. Our response was necessarily iterative, pragmatic, and often pre-dated formal recommendations from state and federal government. During these outbreaks, a number of key factors emerged that shaped our responses, extending well beyond a focus on PPE alone.

First, the concept of a ‘critical burden’ of infection framed our responses to patient movement and ward closures. Concurrent with large numbers of cases in the hospital and the community, the number of staff who acquired infection rose rapidly. Based on overseas experience (9, 10), we hypothesised that large numbers of patients in a confined space may have created a higher density of droplets or aerosolised particles and environmental contamination that may have been important. This concept triggered a detailed assessment of the physical layout of each ward, including the possible role of patient placement and air circulation. RMH was built in 1944 with progressive additions over time, and thus different ventilation systems exist depending on the age of each ward and building. The intensity of transmission in these wards led to a decision to close four wards, and to move some patients to other health care services. In addition, reports of staff experiences derived from case interviews also informed our response. For example, several staff at RPC noted that COVID-19 patients who had a delirium and vocalising loudly may have contributed to transmission of infection which led to a decision to cease cohorting patients. Further we adopted the use of N95 respirators for staff working in areas with large numbers of confirmed or suspected COVID-19 patients (COVID wards) as well as areas of the emergency department and ICU. While we acknowledge this was an area of controversy at the time (11, 12), this organisational decision was based on our own emerging local evidence and a need to trial any reasonably available strategy to contain healthcare workers infections.

Second, the availability of rapid and accessible testing for staff was critical to informing real-time outbreak management. In our health service, testing results were generally available within 12-24 hours of sample collection, with easy access for staff to testing facilities. Recent work from overseas highlights the importance of access to testing for healthcare workers (13, 14). Rapid availability of data informed our daily incident management meetings and enabled decisions to be made using the best possible information. Detailed work, including genomic analysis and a case control study, is underway to understand better and assess the specific risk factors for healthcare acquisition of COVID-19 at our health service.

Finally, the importance of staff communication and well-being cannot be understated. Similar to other studies (4, 15), many staff reported physical and mental fatigue and stress during these outbreaks. In addition, workforce shortages meant that staff were taking on extra shifts at short notice and working in unfamiliar roles. Accordingly, access to employee support programmes was an important element of this response.

Although we acknowledge the descriptive nature of our study, and the lack of rigorous evidence to support aspects of our response, we believe our real-world experience has several learning points for other institutions across Australia. Further work should include improved state and national coordination regarding organisational responses to healthcare worker infections.

## Data Availability

Data are the property of the health service

## Acknowledgments

Chris Baillie, Vivian Leung and Neha Verma for support in data collection and analysis.

## REFERENCES

1. WHO coronavirus disease (COVID-19) dashboard. Geneva: World Health Organization. https://covid19.who.int/ (accessed 2/9/20)

2. Nguyen LH, Drew DA, Graham MS, Joshi AD, Guo CG, Ma W, et al. Risk of COVID-19 among front-line health-care workers and the general community: a prospective cohort study. Lancet Public Health. 2020. Sep;5:e475–e483.

3. Chou R, Dana T, Buckley DI, Selph S, Fu R, Totten AM. Epidemiology of and Risk Factors for Coronavirus Infection in Health Care Workers: A Living Rapid Review. Ann Intern Med. 2020;173:120–36.

4. Bielicki JA, Duval X, Gobat N, Goossens H, Koopmans M, Tacconelli E, et al. Monitoring approaches for health-care workers during the COVID-19 pandemic. The Lancet Infect Dis. 2020. Jul 23;S1473–3099: 30458–8

5. Seemann T, Lane C, Sherry N, Duchene S, Goncalves da Silva A, Caly L, et al. Tracking the COVID-19 pandemic in Australia using genomics. medRxiv. 2020:2020.05.12.20099929.

6. Muhi S, Irving LB, Buising KL. COVID-19 in Australian health care workers: early experience of the Royal Melbourne Hospital emphasises the importance of community acquisition. Med J Aust 2020;213:44–.e1.

7. Williams E, Bond K, Zhang B, Putland M, Williamson DA. Saliva as a non-invasive specimen for detection of SARS-CoV-2. J Clin Microbiol 2020 Jul 23;58(8):e00776–20.

8. Caly L, Druce J, Roberts J, Bond K, Tran T, Kostecki R, et al. Isolation and rapid sharing of the 2019 novel coronavirus (SARS-CoV-2) from the first patient diagnosed with COVID-19 in Australia. Med J Aust 2020 Jun;212(10):459–462

9. Liu Y, Ning Z, Chen Y, Guo M, Liu Y, Gali NK, et al. Aerodynamic analysis of SARS-CoV-2 in two Wuhan hospitals. Nature. 2020;582:557–60.

10. Klompas M, Baker MA, Rhee C. Airborne Transmission of SARS-CoV-2: Theoretical Considerations and Available Evidence. JAMA 2020 Jul 13 doi: 10.1001/jama.2020.12458.

11. Bartoszko JJ, Farooqi MAM, Alhazzani W, Loeb M. Medical masks vs N95 respirators for preventing COVID-19 in healthcare workers: A systematic review and meta-analysis of randomized trials. Influenza Other Respir Viruses. 2020;14:365–73.

12. Dau NQ, Peled H, Lau H, Lyou J, Skinner C. Why N95 Should Be the Standard for All COVID-19 Inpatient Care. Ann Intern Med 2020 Jun 29;M20–2623

13. Keeley AJ, Evans C, Colton H, Ankcorn M, Cope A, State A, et al. Roll-out of SARS-CoV-2 testing for healthcare workers at a large NHS Foundation Trust in the United Kingdom, March 2020. Euro surveillance: bulletin European sur les maladies transmissibles = European communicable disease bulletin. 2020;25(14).

14. Kluytmans-van den Bergh MFQ, Buiting AGM, Pas SD, Bentvelsen RG, van den Bijllaardt W, van Oudheusden AJG, et al. Prevalence and Clinical Presentation of Health Care Workers with Symptoms of Coronavirus Disease 2019 in 2 Dutch Hospitals During an Early Phase of the Pandemic. JAMA Netw Open. 2020;3(5):e209673.

15. Liu Q, Luo D, Haase JE, Guo Q, Wang XQ, Liu S, et al. The experiences of health-care providers during the COVID-19 crisis in China: a qualitative study. Lancet Glob Health. 2020;8(6):e790–e8.

